# Assessing Immunity by Quantitative Measurement of SARS-CoV-2 IgG Antibodies in Fingerstick Samples

**DOI:** 10.1101/2021.12.21.21268075

**Authors:** Javier T. Garza, Jacob Quick, Dev Chatterjee, Robert P. Garr, Atul Varadhachary, Leo Linbeck

**Author notes:** Corresponding Authors: Javier Garza, Dev Chatterjee. These authors contributed equally (Javier T. Garza, Jacob Quick, Dev Chatterjee, Leo Linbeck). Competing Interests: JTG, JQ, DC, AV, and LL are employees of Brevitest Technologies, Inc. Author Contributions: All authors made contributions to the conception, design, or execution of the work. All authors have reviewed the manuscript critically and have given final approval to the manuscript.

## Abstract

COVID-19 has affected billions of people around the world directly or indirectly. The response to the pandemic has focused on preventing the spread of the disease and improving treatment options. Diagnostic technologies have played a key role in this response since the beginning of the pandemic. As vaccines and other treatments have been developed and deployed, interest in understanding and measuring the individual level of immune protection has increased. Historically, use of antibody titers to measure systemic immunity has been constrained by an incomplete understanding of the relationship between antibodies and immunity, the lack of international standards for antibody concentration to enable cross-study comparisons, and insufficient clinical data to allow for the development of robust antibody-immunity models.

However, these constraints have recently shifted. With a deeper understanding of antibodies, the promulgation of WHO antibody standards, and the development of immunity models using datasets from multiple COVID-19 vaccine trials, certain types of quantitative antibody tests may now provide a way to monitor individual or community immunity against COVID-19. Specifically, tests that quantitate the concentration of anti-RBD IgG –antibodies that target the receptor binding domain of the S1 spike protein component of the SARS-CoV-2 virus – show promise as a useful and scalable measure of the COVID-19 immunity of both individuals and communities. However, to fulfill this promise, a rapid and easy-to-administer test is needed.

To address this important clinical need, Brevitest deployed its point-of-care-capable technology platform that can run a rapid (<15 minute), quantitative antibody test with a sample of 10 μl of whole blood from a fingerstick. The test we validated on this platform measures the concentration of anti-RBD IgG in Binding Antibody Units per milliliter (BAU/mL) per WHO Reference Standard NIBSC 20/136.

In this paper, we present studies used to characterize the Brevitest anti-RBD IgG assay and evaluate its clinical performance, lower limits of measurement, precision, linearity, interference, and cross-reactivity. The results demonstrate the ability of this assay to measure a patient’s anti-RBD IgG concentration. This information, together with models developed from recent COVID-19 vaccine clinical trials, can provide a means of assessing the current level of immune protection of an individual or community against COVID-19 infection.

## Introduction

Severe Acute Respiratory Coronavirus 2 (SARS-CoV-2) is a novel coronavirus that emerged as the cause of severe respiratory disease (COVID-19) in December 2019 in China and quickly spread, causing a worldwide pandemic. The SARS-CoV-2 virus is a part of the Betacoronavirus genus of the Coronaviridae family. Other members include the Severe Acute Respiratory Syndrome Coronavirus 1 (SARS-CoV-1) and Middle East Respiratory Syndrome Coronavirus (MERS-CoV).^1^ The high transmissibility of the virus led to its rapid, worldwide spread despite unprecedented efforts by public health authorities to slow the pandemic’s progress, including lockdowns, mask mandates, public hygiene promotions, public investments in rapid vaccine and therapeutic development, and other measures.

Diagnostic testing for detection of active infection by the virus and assessment of the body’s immune response to a viral challenge have played critical roles in efforts to respond to the pandemic, as well as to understand the disease and its effects on the body. For direct detection of the virus in the airways, a plethora of nucleic acid amplification assays (including PCR assays) and viral antigen assays have been developed. These tests have proven to be important elements of clinical diagnosis and disease management strategies. However, viral detection and quantitation assays do not provide information about the body’s immune history, response, or status, nor do they give any indication of the risk of future infection or re-infection.

To understand how a patient’s immune system responds to the virus, we need serological assays. Prior to widespread molecular testing and vaccination, these assays were sometimes used to also diagnose infection by SARS-CoV-2. The rationale for this approach was that the presence of antibodies against a novel virus, especially IgM antibodies, without prior history of infection could be used to diagnose current infection if coupled with concordant clinical symptoms. However, this diagnostic approach was largely abandoned early in the pandemic with the rapid growth and easy availability of antigen and nucleic acid testing (NATs). Nevertheless, serological testing still found some uses for epidemiological monitoring and outbreak control in certain populations.

As the pandemic progressed, collaboration between governments, academics, and the pharmaceutical industry led to the development and testing of several vaccine candidates, culminating in a series of approvals by health authorities worldwide beginning in late 2020. While vaccination provided a significant impact on the extent and severity of infections in most cases, an inevitable decay in that protection was expected based upon prior experience with vaccines. However, the character of that decay and the effective duration of protection varies widely between vaccines and individuals, and the community impact of the loss of protection remains unknown. Accordingly, there is an ongoing debate about the utility, need and timing of booster doses of vaccine to maintain immune protection against the virus, a debate that has intensified with the emergence of different variants of the original strain. To understand and accurately portray an individual’s response to vaccination to help them make individualized decisions with the best available knowledge, quantitative antibody tests are needed that can sensitively measure the antibody levels generated by the vaccine, natural infection, or a combination of the two. But while sensitivity, specificity, and repeatability are essential, rapid results and ease of administration (including both sample matrix and testing location) are also desirable characteristics of any test that is expected to be widely used.

Multiple manufacturers have created and marketed serological assays. The technology behind most of these immunoassays could be categorized as either lateral flow tests (LFT) or enzyme linked immunoassays (ELISA). LFTs are simple to use and require a small amount of blood but most accurate LFTs require a venipuncture to obtain a serum or plasma sample for the test. More recently, a few serological LFTs have been developed that can use fingerstick blood samples. Examples of commercially available LFTs are InBios’ SCoV-2 Detect IgG Rapid Test^2^ and Access Bio’s CareStart COVID-19 IgM/IgG^3^. Despite their ease of use and rapid turnaround of results, a challenge of LFTs is their lack of quantitation and inherently lower sensitivity compared to laboratory ELISAs^4^. The sensitivity limitation is due to the absence of enzyme amplification of the signal, and the lack of washing steps between reactions to separate the reactant chemicals. Nonetheless, LFTs play a significant role in diagnostics because of their low cost, speed, and ease of use.

Another major category of immunoassay tests are ELISAs. Enzyme linked immunosorbent assays are usually performed using one of two methods:

1. ELISA lab kits meant to be run in certified laboratories using 96-well microplates in conjunction with plate readers (e.g., the ZEUS ELISA SARS-CoV-2 IgG Test System^5^; or the EUROIMMUN Anti-SARS-CoV-2 ELISA (IgG)^6^).
2. Preloaded cassettes meant to be run on proprietary laboratory immunoanalyzers (e.g., Roche’s Elecsys Anti-SARS-CoV-2^7^ ; or Siemen’s ADVIA Centaur SARS-CoV-2 IgG (sCOVG)^8^). These ELISAs are higher sensitivity tests which are often semi-quantitative in nature.

Unfortunately, these methods lack the speed and ease of use of LFTs, requiring venipuncture blood as the clinical sample and trained personnel to run the tests in a certified laboratory, which often must make significant capital investments to achieve high levels of testing capacity. Turnaround of results are usually in a few days, depending on the efficiency of the testing operation. However, even the most efficient operations can only deliver test results within a few hours given the limitations of the underlying immunoassay technology.

Brevitest Technologies, Inc. (Brevitest) has developed an immunoassay platform that performs enzyme linked immunoassays at the point of care within 15 minutes. It combines the benefits of LFT (simplicity, low sample volume, cost, and speed) and ELISA (sensitivity, specificity, precision, quantitation, and reliability). The Brevitest platform consists of a sample processing unit (SPU) that runs individual tests on disposable test cartridges. The SPU is a small robotic device that is fully automated and interacts with a central analysis and database system that runs in the cloud (the Brevitest Cloud). It uses a proprietary cartridge that contains reagents required to perform all ELISA steps and uses heat and moving magnets to control the reaction process. When the reactions are complete, the SPU measures the change in color in the indicator reagent resulting from the last step of the ELISA process. The raw optical readings are securely transmitted to the Brevitest Cloud for analysis and processing before the result is made available to the patient and physician in real-time. We have previously published the use of this platform to detect the presence of anti-SARS-CoV-2 secretory IgA in saliva.^9^

Brevitest has developed and validated an assay to quantify SARS-CoV-2 anti-RBD IgG antibodies in fingerstick blood in less than 15 minutes on its proprietary platform. In this report, we present data from the validation studies and discuss the test’s potential clinical and epidemiological uses.

## Materials and Methods

### Materials, instruments, and reagents

The Brevitest assay platform has three components: a sample processing unit (SPU), a pre-loaded cartridge, and cloud-based analytical software (Brevitest Cloud). (See Figure 1.) The SPU is a miniaturized fully automated robotic device with mechanical, optical, and thermoregulatory elements. The SPU uses permanent magnets to control the movement of magnetic microspheres across several compartments within the cartridge and oscillates the magnets in each compartment using proprietary mixing patterns. In each compartment, a single step of the multi-step ELISA reaction takes place on the surface of these microspheres. The final reaction results in production of a colored compound that is measured as an optical signal, and the raw readings are sent to the Brevitest Cloud for analysis. There are several safeguards built into the platform to ensure that the cartridge is inserted properly, the correct test is performed, the cartridge is not past its expiry date, test results are valid, and patient identifiable information is not at risk.

**Figure 1.**
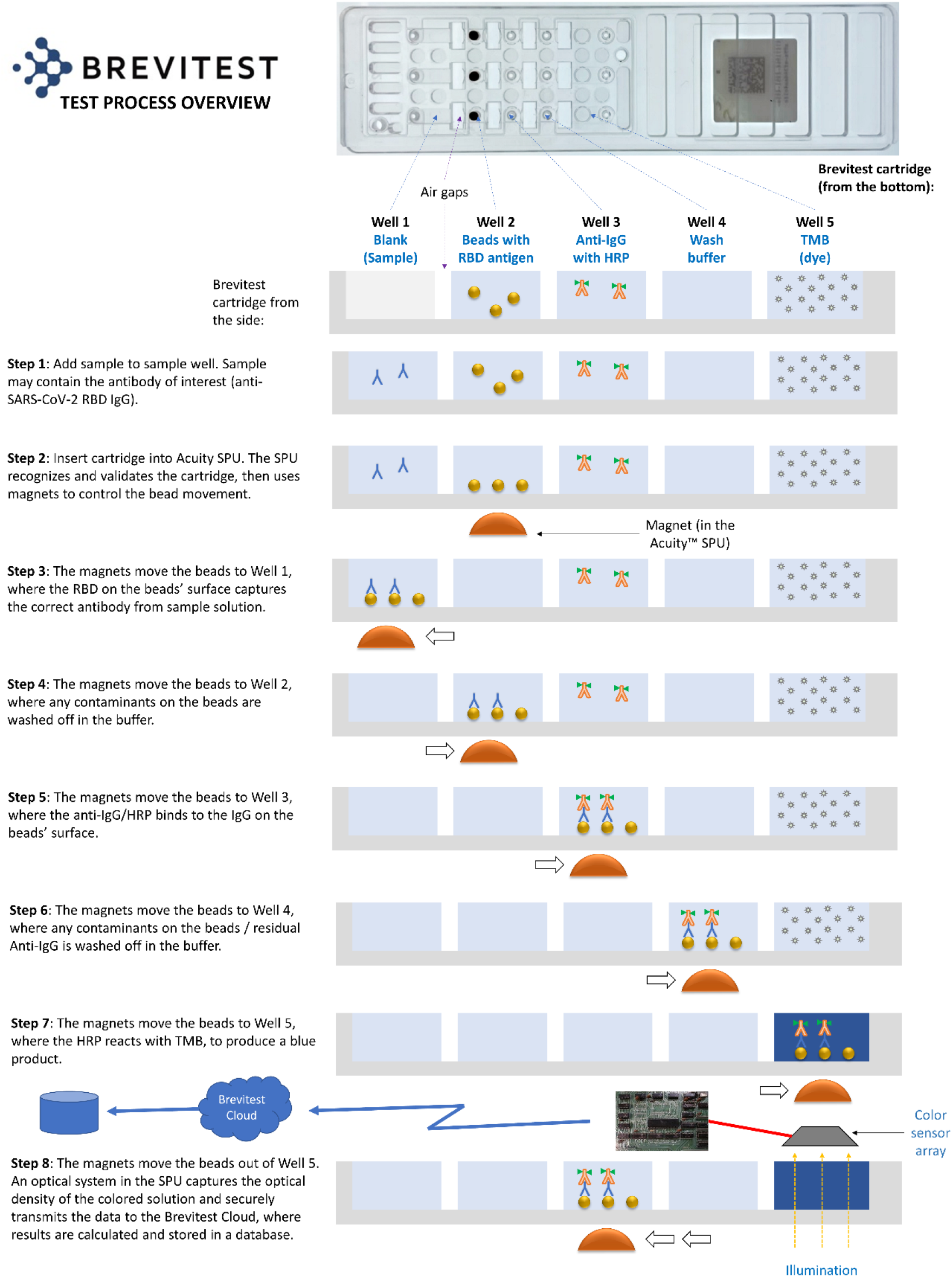
Brevitest Test Process Overview

The cartridge is pre-loaded with the reagents required to run Brevitest’s COVID-19 IgG assay. The cartridge is not a microfluidic device, as there is no movement of reagents within the cartridge; rather, the key ELISA components – antigen, capture antibody, and tracer antibody – are bound to magnetic microspheres that are moved between reagent wells. The capture antigen used is the Receptor Binding Domain (RBD) of the S1 protein from the SARS-CoV-2 virus. The captured IgG is then detected using an anti-human IgG linked to a horseradish peroxidase (HRP) molecule. The final color change occurs in the compartment containing TMB, which changes from colorless to blue in the presence of HRP. Each cartridge is configured to run 3 ELISA reactions simultaneously in separate channels. Two channels contain the sample at high and low dilution, respectively, and one channel is loaded with a known concentration of anti-SARS-CoV-2 Spike S1 IgG for quality control. The testing materials, which include the SPU and the cartridges, were developed at Brevitest Technologies, Inc. and manufactured at their CLIA-certified laboratory.

Samples were collected from subjects according to an IRB-approved protocol after consent with an IRB-approved informed consent form. Samples include fingerstick blood obtained according to standard procedures with appropriate safety precautions. To collect the fingerstick blood for the Brevitest assay, a 28G disposable lancet was used, followed by collection with a 10 μl disposable fixed volume microcapillary tube. The manufacturer’s data for the microcapillary tube indicate that sample volume varies <1% on clinical tests. The clinical history of the subjects was collected in a questionnaire and recorded before taking their sample.

### Clinical Validation

Fingerstick samples (107 samples) were obtained from individuals who responded to a call to participate in the clinical study conducted at Brevitest’s CLIA-certified laboratory in Houston, Texas. Ninety (90) samples were obtained from those with a reported history of SARS-CoV-2 infection and/or those who had received at least one dose of any of the three vaccines, and were classified as “positive.” Seventeen (17) samples were obtained from individuals who did not have a medical history of infection with SARS-CoV-2 and reported as never receiving a COVID-19 vaccine, and were classified as “negative.”

### Precision

The precision of the assay was characterized in a study guided by CLSI EP05-A3. In this study, three samples representative of the spectrum of concentrations encountered in routine clinical operations were tested. The samples were tested in replicates of two for three days on two different lots per day (for a total of six lots) and three analyzers. The software *Analyse-it* was used to calculate the results.

### Cross-reactivity and Interference

To test for cross-reactivity on the Brevitest assay, a panel of samples known to be negative for SARS-CoV-2 and positive for IgG antibodies to the following viruses were obtained from BioIVT: influenza A, influenza B, HBV, HCV, HIV, Haemophilus influenzae, alpha coronaviruses 229E and NL63, beta coronaviruses OC43 and HKU1, RSV, Parainfluenza, and Mycoplasma pneumoniae. These samples were tested on the Brevitest assay, and the ones with results below the limit of detection (LOD; 6.2 BAU/mL) were classified as non-cross reactive.

Common interferents were acquired from Sun Diagnostics to spike samples and determine their effect on the test. Four fingerstick samples not containing the interferent (low and high SARS-CoV-2 IgG samples) were spiked with hemolysate (1000 mg/dL), triglyceride-rich lipoproteins (500 mg/dL), and conjugated bilirubin (40 mg/dL). The percent difference to the control sample was calculated for each spiked sample and the results interpretations were examined.

### Assay Analytical Performance

The detection capabilities of the assay, including the limit of blank (LOB) and LOD, were determined based on guidance from CLSI EP17-A2. To obtain the LOB, 4 blank fingerstick samples were tested in replicates of 2 for 3 days on 2 lots and 3 analyzers. LOD was determined by testing 4 low level samples in replicates of 2 for 3 days on 2 lots and 3 analyzers. The 4 low level samples were created by mixing confirmed negative samples with a known amount of WHO standard (0.17 μl of 1000 BAU/mL standard). The limit of quantitation (LOQ) was calculated as lowest concentration tested greater than the LOD plus 4.5 times the standard deviation observed in our LOD study. The statistical analysis was conducted with the software *Analyse-it*. A study guided by CLSI EP06-A was performed to determine the linearity. Blank samples were spiked with the international WHO standard (WHO Reference Standard NIBSC 20/136) to prepare 10 samples of known concentration. The samples were tested in 4 replicates using 1 reagent lot.

## Results

### Demographic data

A breakdown of the demographics of the tested population is presented below (Table 1).

**Table 1:** Demographic data of test population

|  |  | Positive* | Negative** |
| --- | --- | --- | --- |
| Total |  | 90 | 17 |
| Sex | Male | 49 | 4 |
|  | Female | 40 | 13 |
|  | Unknown | 1 |  |
| Infection | Total | 22 | 0 |
|  | No vaccination | 8 |  |
|  | Also vaccinated | 14 |  |
| Vaccinated | Total | 82 | 0 |
|  | Pfizer | 40 | 0 |
|  | Moderna | 34 | 0 |
|  | J&J | 7 | 0 |
|  | Unknown | 1 |  |
| Time since infection (days) | Median | 263 |  |
|  | Range | 26 – 485 |  |
| Time since last vaccination dose (days) | Median | 155 |  |
|  | Range | 4 - 267 |  |
| * Positive defined as somebody who has a history of COVID-19 infection or vaccination or both.<br>** Negative defined as people without a history of either infection or vaccination. |  |  |  |

### Clinical performance

Due to the impracticality of getting pre-COVID samples of fingerstick whole blood and the lack of a fingerstick reference test for quantitative measurement of COVID-19 IgG antibodies, a clinical agreement study was performed based upon clinical history. Samples were collected from a total of 107 subjects. Clinical history data was also obtained from these subjects; all data was self-reported and verification of data was performed where practical.

Based on this self-reported clinical history data, 17 subjects never had symptomatic infection with COVID-19 and were never vaccinated. The other 90 subjects had either been infected, vaccinated, or both. Subjects that had been neither vaccinated nor infected were expected to have no detectable antibodies (i.e., below the limit of detection) and were characterized as negative. The remaining subjects were expected to have detectable levels of antibodies (i.e. above the limit of detection) and were characterized as positive.

The results showed overall positive percent agreement (PPA)/sensitivity of 97.8% (95% CI: 92.42% - 99.40%) and overall negative percent agreement (NPA)/specificity of 100% (95% CI: 79.61% - 100%). See Table 2.

**Table 2:** Clinical Performance Study Results

|  |  | Reference method |  |  | Performance |  | 95% CI |
| --- | --- | --- | --- | --- | --- | --- | --- |
|  |  | Positive | Negative | Total |  |  |  |
| **Brevitest test results** | Positive | 90 | 2 | 92 | PPV | 97.8% | 92.4-99.4% |
|  | Negative | 0 | 15 | 15 | NPV | 100.0% | 79.6-100.0% |
|  | Total | 90 | 17 | 107 |  |  |  |

### Assay characteristics

To determine the repeatability, within-lot precision, and reproducibility, 3 samples were tested multiple times to obtain the results presented in Table 3.

**Table 3:** Precision Study Results

| Sample | Mean | Repeatability SD | Repeatability CV | Within Lot SD | Within Lot CV | Reproducibility SD | Reproducibility CV |
| --- | --- | --- | --- | --- | --- | --- | --- |
| P1 | 71.9 | 5.7 | 8.0% | 8.7 | 12.0% | 16.8 | 23.4% |
| P2 | 692.5 | 43.8 | 6.3% | 89.9 | 13.0% | 120.6 | 17.4% |
| P3 | 1262.6 | 90.5 | 7.2% | 173.7 | 13.8% | 175.1 | 13.9% |

Several blank and low-level samples were tested to calculate the LOB, LOD, and LOQ. The results, shown in Table 4, present the obtained lower limits of the assay.

**Table 4:** Analytical Sensitivity Study Results

|  | BAU/mL |
| --- | --- |
| LOB | 0.23 |
| LOD | 6.20 |
| LOQ | 22.2 |

In the linearity study, solutions of known concentrations were prepared and tested on the Brevitest device to compare the values obtained. Linearity was demonstrated across the analytical measuring interval of 22.2 to 2000 BAU/mL (see Figure 2). From the 10 samples tested, 2 points, 0 BAU/mL and 4000 BAU/mL, were removed from the linear range (see Table 5). Taking into consideration the estimates of LOB, LOD, LOQ, and linearity, the analytical measuring interval is from 22.2 to 2000 BAU/mL.

**Figure 2.**
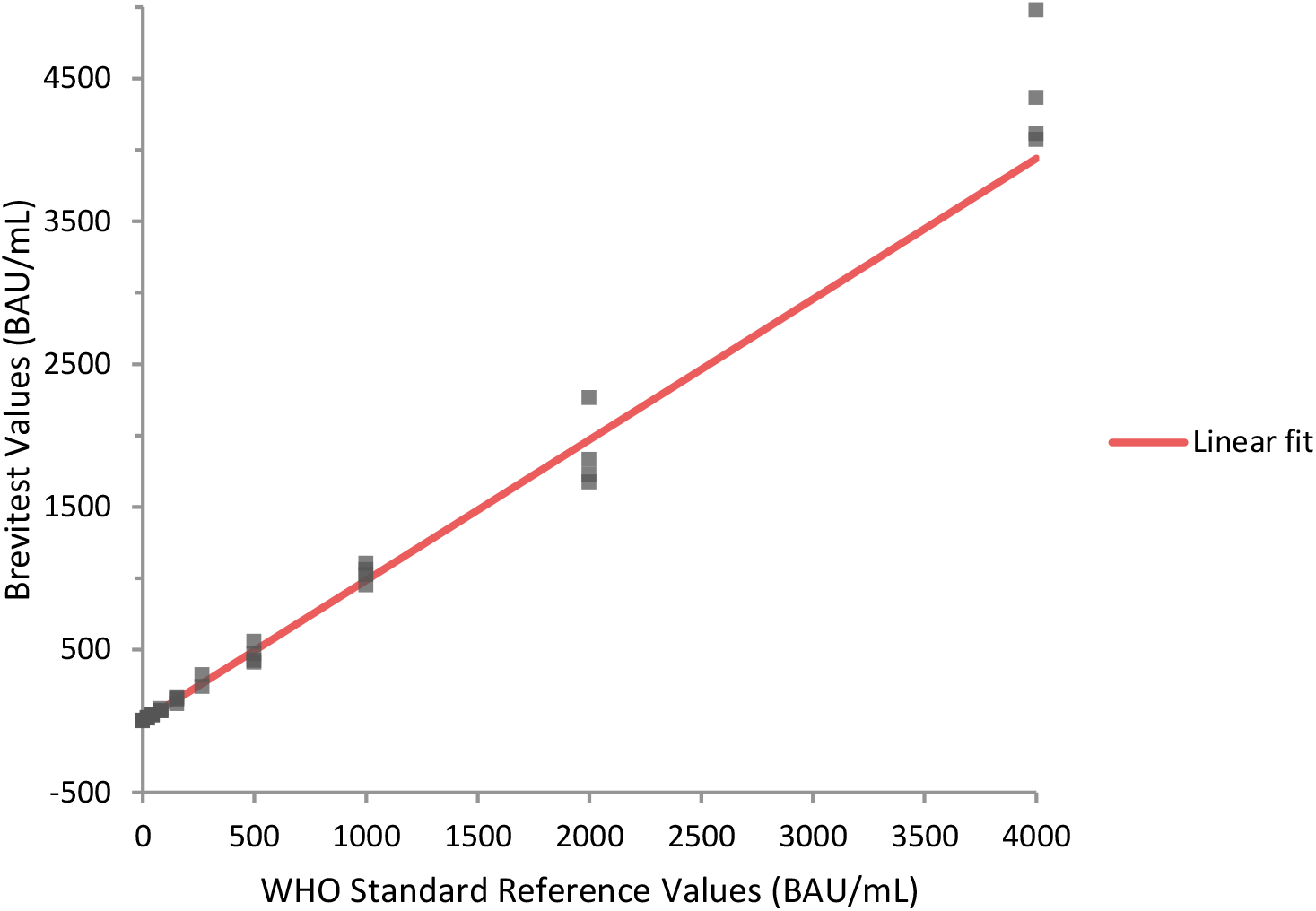
Linearity plot that presents the Brevitest results for each of the 10 samples tested. The samples were prepared with the WHO International Standard. The linear range was determined to be from 22.2 to 2000 BAU/mL.

**Table 5:** Linearity Study Results

| Known Values | Mean | Linear fit | Nonlinearity | Allowable nonlinearity |
| --- | --- | --- | --- | --- |
| 0.00 | 3.20 | 1.38 | 130.9%* | ±10.0% |
| 22.22 | 21.85 | 23.27 | -6.1% | ±10.0% |
| 43.48 | 44.33 | 44.20 | 0.3% | ±10.0% |
| 83.33 | 76.20 | 83.45 | -8.7% | ±10.0% |
| 153.85 | 151.24 | 152.90 | -1.1% | ±10.0% |
| 266.67 | 282.91 | 264.00 | 7.2% | ±10.0% |
| 500.00 | 466.28 | 493.78 | -5.6% | ±10.0% |
| 1000.00 | 1035.72 | 986.18 | 5.0% | ±10.0% |
| 2000.00 | 1874.59 | 1970.97 | -4.9% | ±10.0% |
| 4000.00 | 4383.45 | 3940.56 | 11.2%* | ±10.0% |
| *Performance requirement not met. |  |  |  |  |

### Cross reactivity and interference

We evaluated the assay for potentially cross-reacting antibodies. Some HCV samples (2), HIV samples (1), and all the Coronavirus samples were above the limit of detection for the SARS-CoV-2 IgG test. The rest of the samples were below the LOD (see Table 6).

**Table 6:** Cross-reactivity Study Results

| Category | N | <6.2<br>BAU/mL | 6.2-22.2<br>BAU/mL | ≥22.2<br>BAU/mL |
| --- | --- | --- | --- | --- |
| Human serum Hepatitis B (HBV) | 4 | 4 | 0 | 0 |
| Human serum Hepatitis C (HCV) | 5 | 3 | 1 | 1 |
| Human serum HIV | 5 | 4 | 1 | 0 |
| Coronavirus HKU | 5 | 0 | 2 | 3 |
| Coronavirus OC43 | 5 | 0 | 2 | 3 |
| Coronavirus 229E | 5 | 0 | 2 | 3 |
| Coronavirus NL63 | 5 | 0 | 2 | 3 |
| H. Influenzae | 2 | 2 | 0 | 0 |
| RSV | 2 | 2 | 0 | 0 |
| Influenza B IgG | 2 | 2 | 0 | 0 |
| Parainfluenza 1-4 IgG | 2 | 2 | 0 | 0 |
| M. pneumoniae IgG | 2 | 2 | 0 | 0 |

We also evaluated potentially interfering substances (see Table 7). Each substance was tested in 4 samples with different levels of SARS-CoV-2 IgG (low and high). After spiking the samples with the interferents, the results were analyzed by comparing them with the samples without the interferents. The percent difference was calculated for each sample. All the samples, except the lowest sample spiked with hemolysate, were less than 15% percent difference. The results interpretations did not change for any of the samples. Table 8 summarizes the results obtained.

**Table 7:**
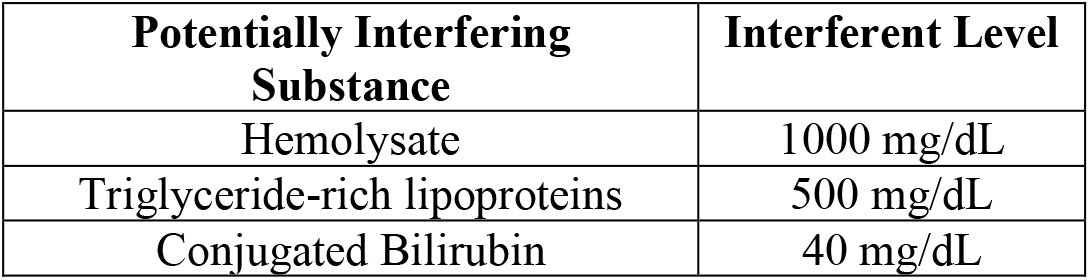
Interfering Substances Tested

**Table 8:**
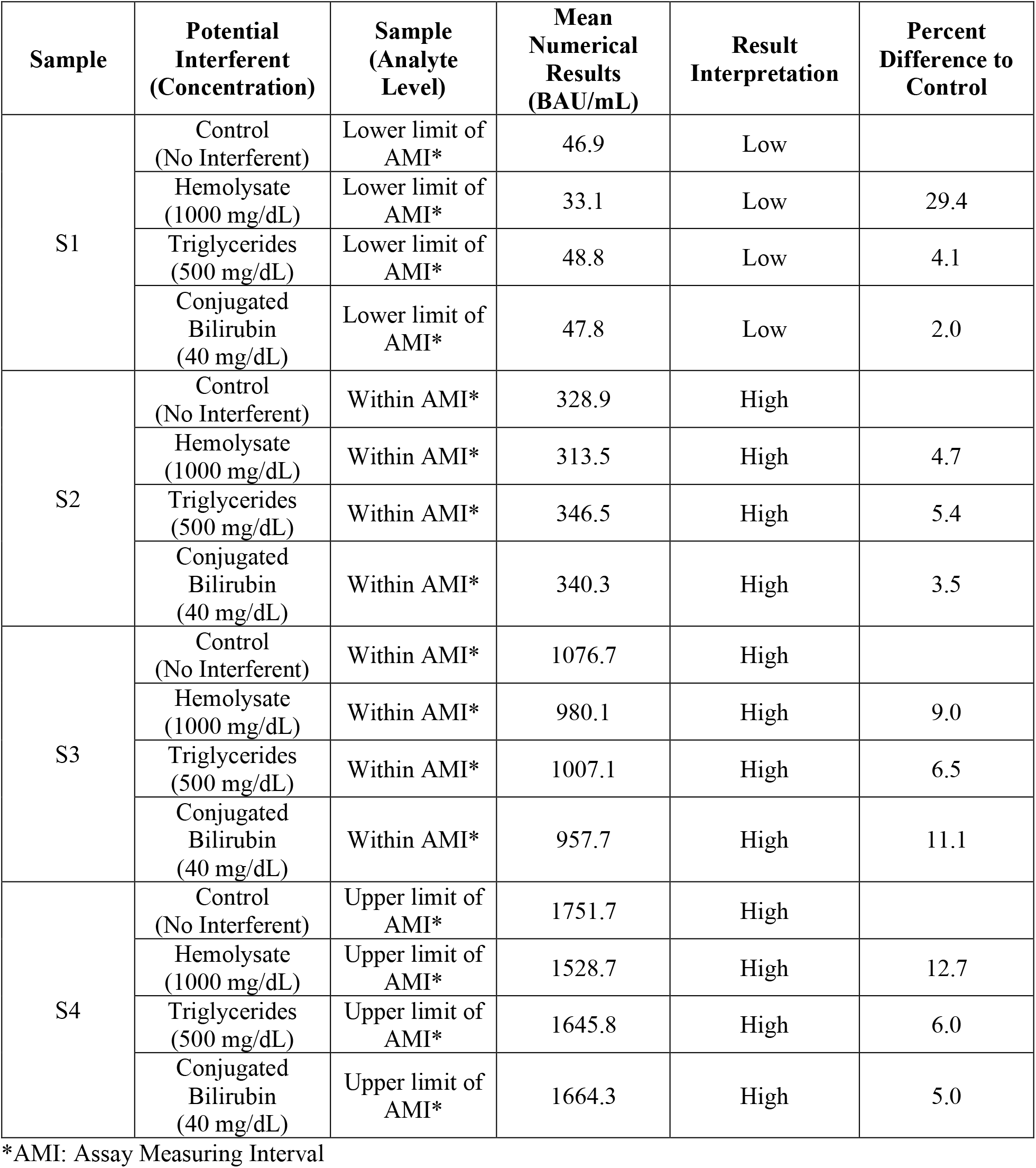
Interference Study Results

| Sample | Potential Interferent (Concentration) | Sample (Analyte Level) | Mean Numerical Results (BAU/mL) | Result Interpretation | Percent Difference to Control |
| --- | --- | --- | --- | --- | --- |
| S1 | Control (No Interferent) | Lower limit of AMI* | 46.9 | Low |  |
|  | Hemolysate (1000 mg/dL) | Lower limit of AMI* | 33.1 | Low | 29.4 |
|  | Triglycerides (500 mg/dL) | Lower limit of AMI* | 48.8 | Low | 4.1 |
|  | Conjugated Bilirubin (40 mg/dL) | Lower limit of AMI* | 47.8 | Low | 2.0 |
| S2 | Control (No Interferent) | Within AMI* | 328.9 | High |  |
|  | Hemolysate (1000 mg/dL) | Within AMI* | 313.5 | High | 4.7 |
|  | Triglycerides (500 mg/dL) | Within AMI* | 346.5 | High | 5.4 |
|  | Conjugated Bilirubin (40 mg/dL) | Within AMI* | 340.3 | High | 3.5 |
| S3 | Control (No Interferent) | Within AMI* | 1076.7 | High |  |
|  | Hemolysate (1000 mg/dL) | Within AMI* | 980.1 | High | 9.0 |
|  | Triglycerides (500 mg/dL) | Within AMI* | 1007.1 | High | 6.5 |
|  | Conjugated Bilirubin (40 mg/dL) | Within AMI* | 957.7 | High | 11.1 |
| S4 | Control (No Interferent) | Upper limit of AMI* | 1751.7 | High |  |
|  | Hemolysate (1000 mg/dL) | Upper limit of AMI* | 1528.7 | High | 12.7 |
|  | Triglycerides (500 mg/dL) | Upper limit of AMI* | 1645.8 | High | 6.0 |
|  | Conjugated Bilirubin (40 mg/dL) | Upper limit of AMI* | 1664.3 | High | 5.0 |

\*AMI: Assay Measuring Interval

## Discussion

It is often said that infection is a war between pathogen and host. In that war, the central element of our defense – whether against an individual infection or a global pandemic – is the human immune system.

At its most basic level, the immune system is composed of cells, proteins, and other molecules that work in concert to identify pathogens, prevent infection and, should an infection take hold, eliminate the invading pathogen or reach a sustainable “stalemate” with it.

Continuing with the war analogy, the immune system has an extensive “intelligence” capability that uses cell surface receptors, cytokines, chemokines, dendritic cells, immunoglobulins, major histamine complexes, and other elements to recognize toxins, bacteria, fungi, parasites, and viruses, and send alerts to other parts of the immune system that a threat has been detected and action is required.

In response, the immune system mobilizes a wide range of “weaponry” to neutralize and eliminate the threat, including, but not limited to reactive oxygen species, anti-microbial peptides, effector B cells, plasma cells, CD4 T cells, CD8 T cells, NK cells, macrophages, neutrophils, and an extraordinarily diverse range of free immunoglobulins, or antibodies. These components can be viewed as elements of three major subsystems – innate immunity, mucosal immunity, and humoral immunity – but the interconnectedness and interaction of these subsystems is extensive, adding both redundancy and complexity to the overall immune system.^10^

Antibodies are important effectors of both mucosal and humoral immunity, but also participate in pathogen recognition and signaling processes. Their two primary effector modes are neutralization and opsonization, with neutralization being of especial importance when challenged with a viral pathogen. Vaccine researchers and developers have paid particular attention to neutralizing antibodies because of their ability to prevent entry of viral RNA into host cells, thus impeding the viral replication process that is a necessary component of any serious infection.

The protective role played by antibodies has been demonstrated in COVID-19 with both convalescent serum and with therapeutic monoclonal antibodies, which have proved to be especially beneficial in individuals with low levels of endogenous antibodies.^11,12^ FDA has approved monoclonal antibodies therapeutically in individuals recently diagnosed with COVID-19 and prophylactically in individuals unable to mount an immune response against SARS-CoV-2.^13^

However, while antibodies play important biological roles as *effectors of immunity*, they could have even more important clinical and epidemiological roles as *indicators of immunity*.

These roles are often confounded; we regularly hear that “immunity is more than antibodies” as a reason to not test for antibody levels. This observation, of course, is true. It takes more than antibodies to generate the sort of robust, redundant, and rapid response required for the survival of the human species in the hostile environment in which we live.

But the question is not whether antibody levels are *synonymous* with immunity; the question is whether antibody levels are *correlated* with immunity.

From a clinical perspective, the most important measure of overall immune protection is reduction in the risk of symptomatic infection compared to the general population. (Asymptomatic infections may be relevant to certain epidemiological questions, but they are not typically considered a disease state of clinical importance.) This metric has been widely used for assessing the efficacy of a COVID-19 vaccines.^14^ Vaccine trials, for instance, have a primary endpoint of “vaccine effectiveness,” which is the reduction of the rate of infection in the active (vaccinated) arm relative to the control (placebo) arm.

However, because infection rates are usually quite low, vaccine trials by necessity have very large numbers of subjects. The duration and cost (both in money and risk to human health) of such trials have been a matter of significant concern to policymakers and industry, as they constrain responses to fast-moving viral outbreaks that can become global pandemics. As a result, there has long been a desire to find a surrogate biomarker that can be used for assessing the immune status of individuals and populations, and thus guide both vaccine development and pandemic response management.

In short, we want an easy way to measure immunity.

Given what we know about how the immune system works, antibodies are an obvious category of measurands to consider for this role. They are involved in both the early response to a pathogen challenge (through generation of pentameric IgM and dimeric secretory IgA) and longer-term immunity through memory B cells that differentiate into antibody-producing plasma cells when a pathogen reappears and generates a secondary immune response.^15^

However, the use of antibodies as correlates of immunity has been hindered by the lack of answers to two fundamental questions:

1. *What should we measure?* The diversity of antibody classes and targets makes the selection of the specific antibody to measure very difficult. Even if the use case makes class determination somewhat easy (e.g., focus on IgG for vaccine development), the question of target antigen remains challenging. In the case of SARS-CoV-2, for instance, do we want antibodies that bind to a specific viral component (e.g., the spike protein)? Given a component, do we want to bind to any segment or a specific one (e.g., S1)? Given a segment, do we want to bind to any domain or a specific one (e.g., RBD)? And what happens when mutations occur, causing a change in the potential binding sites?
2. *How should we measure?* Most antibody assays used today are simple lateral flow tests that give a binary result (+/-). LFTs are widely administered because they are easy to use (some requiring only a fingerstick), but they lack the sensitivity and precision needed to provide clinicians with a tool for determining immune status. There are semi-quantitative assays that can provide better information, but they typically require large samples of venipuncture blood, thus making widespread adoption difficult and unattractive to many patients. And, finally, there has until recently not been an international standard for antibody units, so semi-quantitative tests were of little utility when trying to assess community-wide immunity.

The recent emergence of an international standard for antibody concentration was an important first step in answering these two questions. Without such a standard, the comparison of antibody test datasets ranges from difficult to impractical to impossible. The WHO Reference Standard NIBSC 20/136^16^ was established in December 2020 and is a crucial and welcome development that promises to dramatically accelerate our ability to both find and propagate insights of interest to scientists, clinicians, and policymakers.

Another major recent development that has dramatically enhanced the potential to measure immunity is the unprecedented number and scale of vaccine trials run over the past 12 months. As data from these trials have been published and analyzed, two very important insights have emerged:

A. *Neutralizing antibodies are highly correlated with immunity against COVID-19*.^17^ From the beginning of the pandemic, researchers focused on the spike protein of the SARS-CoV-2 virus. That focus was rewarded and reinforced by data from mRNA vaccine trials, which showed that antibodies against the spike protein generated by the vaccine were highly protective.
B. *Certain antibody tests are good correlates for protection against COVID-19*.^18^ Based upon the above insight, it is not surprising that neutralization assays – both live virus and pseudovirus types – are good correlates for immunity against COVID-19 and lessening of the severity of the disease. However, neutralizing antibody assays are difficult (and, in the case of live-virus assays, dangerous) to perform, thus limiting their availability to only a few clinical settings. However, further research has found that binding antibodies against the RBD domain are also strongly correlated with protection against COVID-19, perhaps due the neutralizing effect of competitive antibody binding that interferes with the binding of the spike protein to the ACE-2 receptor.

All these recent developments – the emergence of an international standard, the identification of neutralizing antibodies as excellent correlates of protection, and the extension of that correlation to other, more accessible antibody tests – have created the conditions for a dramatic improvement in the clinical management of COVID-19 using antibody tests as a guide. But to implement this improvement, a testing technology is needed that can deliver quantitative results in WHO standard units of BAU/mL with sufficient precision at the point of care from a convenient, patient-friendly sample in a rapid time frame.

The Brevitest Quantimmune™ COVID-19 IgG Antibody Test is a quantitative test that presents the results in BAU/mL, in accordance with WHO Reference Standard NIBSC 20/136 for anti-SARS-CoV-2 antibody. In developing this assay, the LOB, LOD, LOQ, reproducibility, and linearity were measured and calculated. The LOD was calculated to be 6.2 BAU/mL, and the LOQ 22.2 BAU/mL. The studies to determine these values were conducted on fingerstick samples from patients that were not previously infected or vaccinated. Samples from patients were also spiked with low levels of WHO standard to create low levels samples used in the calculations. To test linearity, a negative sample was spiked with known levels of the reference WHO standard. Based on the results, the quantifiable range of the assay was determined to be from 22.2 to 2000 BAU/mL. Precision studies demonstrated a within-lot precision of 13.8% and a reproducibility of 23.4 %CV. Cross-reactivity and interference were also tested to determine if potential samples or substances could impact the test result.

Brevitest measures antibodies against the RBD part of the spike protein, which have been shown to be highly correlative with neutralizing activity.^19^ In a recently published study, researchers analyzed the association between the concentration of certain antibodies and protection against SARS-CoV-2 quantified as vaccine effectiveness, that is the reduction in the rate of symptomatic infection for vaccinated vs. unvaccinated subjects.^18^ In the study, binding and neutralizing antibodies at 28 days after the second dose were measured in infected and uninfected vaccine recipients, with antibody concentrations reported using the WHO reference standard. A vaccine effectiveness of 50% against symptomatic infection with majority Alpha (B.1.1.7) variant of SARS-CoV-2 was achieved with 95% confidence (VE50@95) at a concentration of anti-RBD antibodies of 109 BAU/mL.^18^

In the Brevitest assay, this number was used to set the cutoff difference between “Low” and “High,” with values below the LOD reported as “None Detected.” Figure 3 shows the cutoffs of the Brevitest assay. Thus, the results of the Brevitest Quantimmune™ COVID-19 IgG Antibody Test can be used to estimate the level of protection in a clinical setting.

**Figure 3.**
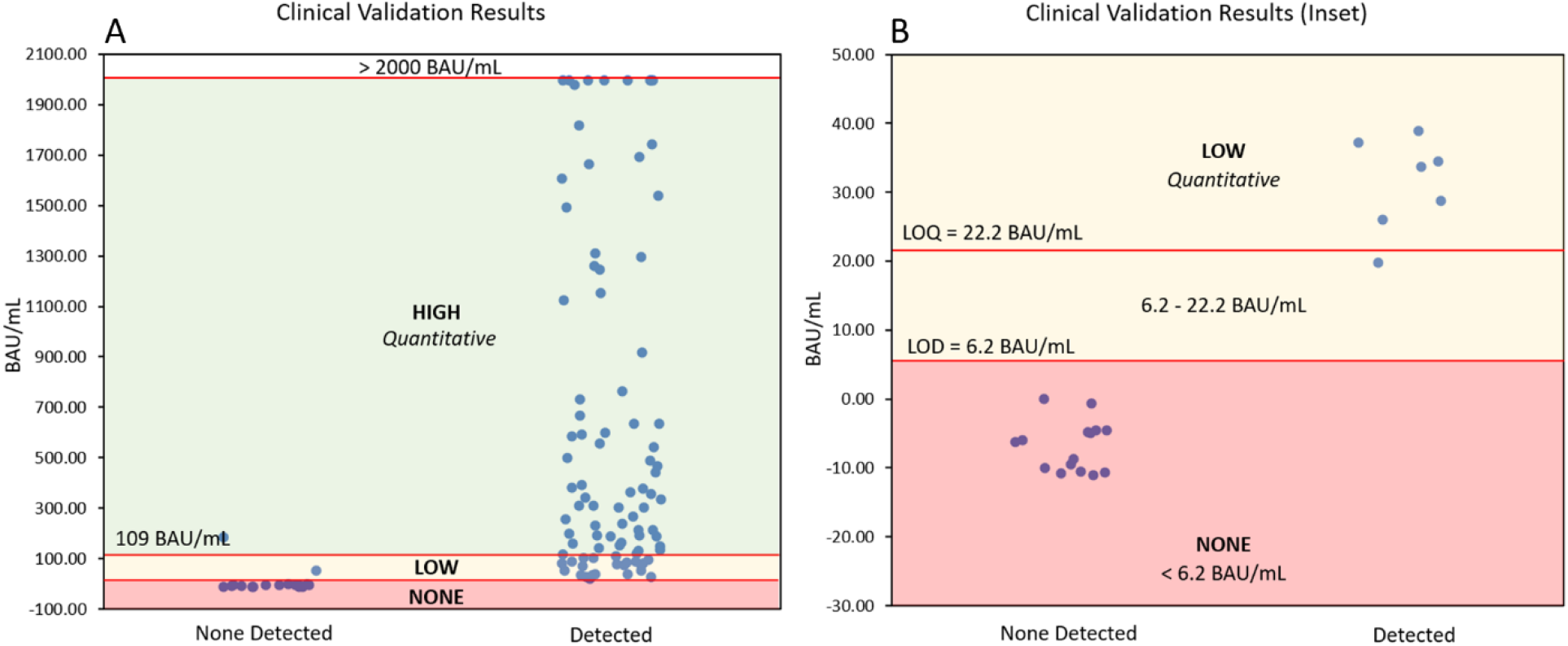
Brevitest assay cutoffs. In Figure 3A, the cutoff of 109 BAU/mL differentiates a low and a high sample. Figure 3B presents the LOD (6.2 BAU/mL) and LOQ (22.2 BAU/mL) cutoffs. A quantitative result is reported from 22.2 to 2000 BAU/mL

The availability of rapid, quantitative serology in a point-of-care setting from a fingerstick that can assess the level of immune protection on an individual and community basis is unprecedented and could have profound impact on the response to pandemics such as COVID-19. For instance, in the administration of vaccine boosters, public health officials and vaccine developers have typically recommended a booster after a specific time. Current recommendation for mRNA vaccines, for example, is to get a booster after 6 months. However, this “fixed booster time” approach can be suboptimal for patients (particularly elderly ones) who tend to lose their immunity faster than 6 months. In addition, for many people vaccination against COVID-19 confers 12 months or more of protection. For that segment of the vaccinated population, it could be optimal to defer a booster until their immunity falls below a threshold. Such a deferral could have multiple benefits: it could extend the overall period of protection for that patient, perhaps generate a stronger and more specific secondary response, and allow for better use of limited vaccine supplies.

Figure 4 presents data taken from the Brevitest clinical validation study, showing only subjects who received an mRNA vaccine (Moderna or Pfizer-BioNTech). While the dataset is relatively small (n=73), more than 12% of subjects were below the VE50@95 level of 109 BAU/mL prior to the 6-month recommended booster date, suggesting they could have a protection gap if they were to wait to boost. On the other hand, more than 31% of subjects had antibody levels above VE50@95, suggesting that they still had residual immunity, at least against the B.1.1.7 variant, and might extend their period of immune protection by deferring a booster. While such decisions should always be made in consultation with a clinician, this approach could be a helpful guide for those trying to manage patient immunity.

**Figure 4.**
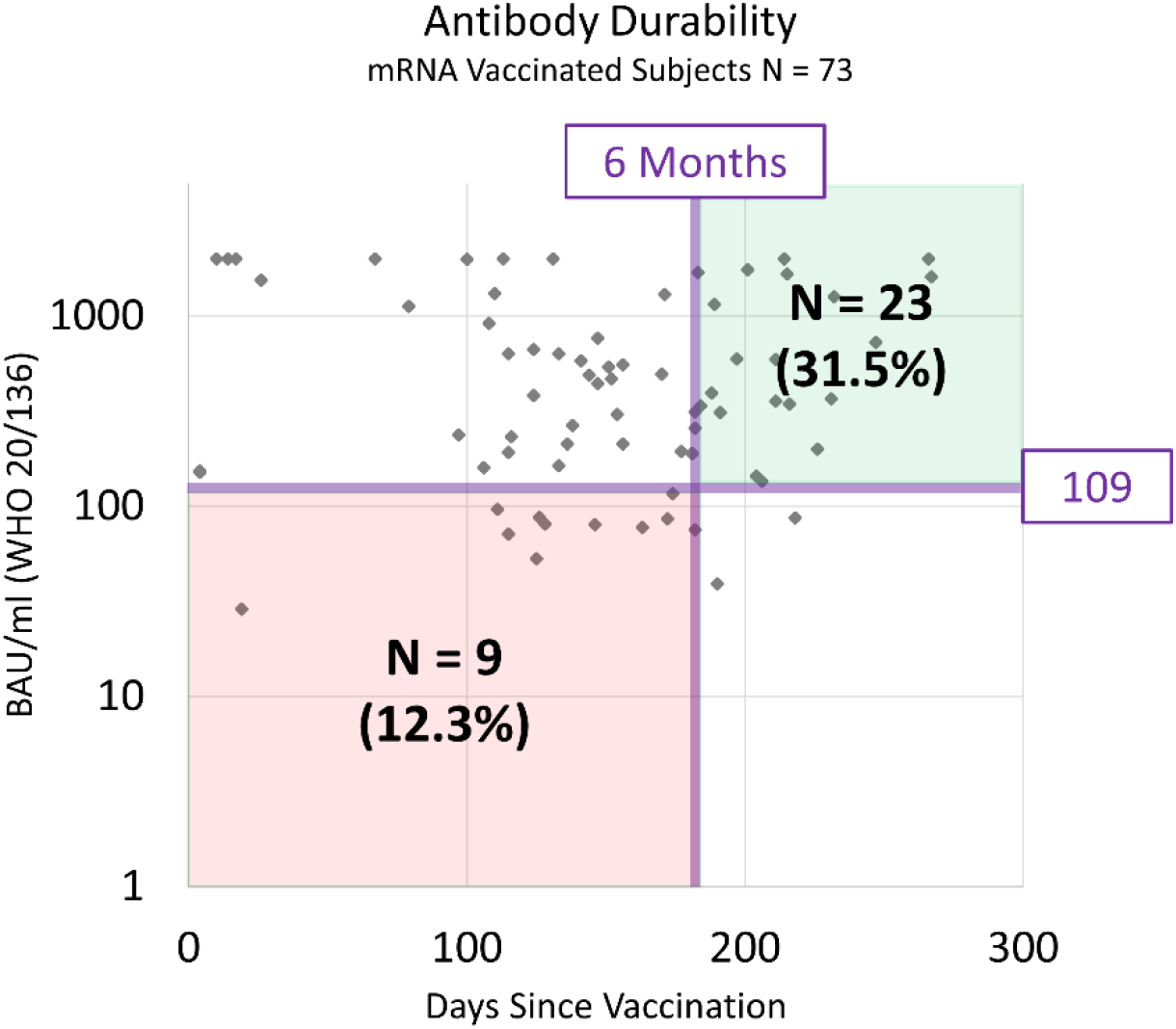
Vertical axis is anti-RBD antibody concentration as measured by the Brevitest Quantimmune™ COVID-19 IgG Antibody Test. Horizontal axis is the days between the self-reported date of the second vaccine dose and the antibody test date. The recommended time after vaccination to receive a booster is 6 months for mRNA vaccines. The VE50@95 level is 109 BAU/mL.

A second clinical use case might be to evaluate the immune response after infection or vaccination. The specifics of seroconversion vary among individuals depending on age, genetic makeup, underlying disease, medication, and other factors. People who get vaccinated produce different types and concentrations of antibodies, and these characteristics change over time. Information about a patient’s current concentration of antibodies can guide lifestyle decisions by factoring in the risks associated with protection levels and possible viral exposure in different environments.

A third use could be in the screening and monitoring of patients undergoing antibody therapy, since decisions can be impacted by the levels of antibody detected. For example, it has been shown that convalescent plasma and therapeutic monoclonal antibody treatments are more effective for patients with no antibodies versus patients that have endogenous antibodies against SARS-CoV-2.^11,12^ Thus, this information can also be used to identify patients that would benefit more from monoclonal antibody administration, either therapeutically or prophylactically, and aid health care providers in managing the administration of those therapies.

Finally, seroprevalence studies could be greatly enhanced by using a quantitative antibody test to assess herd immunity. Whether done at the community level or in certain other appropriate settings such as offices, manufacturing plants, or schools, epidemiological modeling and public health decision-making would be greatly improved with a more accurate, easy-to-administer test, particularly one that is cloud-connected and thus capable of a “bird’s eye” view of immunity by integrating real-time antibody results into a geolocation and geomapping software system.

## Conclusion

Recent studies have shown that a quantitative anti-RBD IgG antibody test is a useful correlate for immune protection against COVID-19. To maximize the impact of this important insight, a test that is rapid (<15 minutes) and precise, requiring only a fingerstick sample of blood, and capable of running in a point-of-care setting is needed.

The Brevitest assay can quantify the amount of SARS-CoV-2 IgG antibodies in fingerstick samples in BAU/mL, per WHO Reference Standard 20/136. Its performance characteristics are presented in this work. In the assay validation tests, the lower limits of measurement, precision, linearity, interference, and cross-reactivity were determined and evaluated.

Among possible uses, this test could be a valuable tool to optimize the timing of a vaccine booster, verify seroconversion in immunocompromised and other at-risk groups, optimize the therapeutic and prophylactic use of monoclonal antibodies and, more broadly, assess individual and population immune responses, specifically production of IgG antibodies, triggered by SARS-CoV-2 or the vaccine. Information about the quantity of SARS-CoV-2 antibodies present in a sample of an individual at a given time could be valuable as it could guide therapeutic, preventative, or lifestyle decisions that could prevent or manage COVID-19.

## Data Availability

All data produced in the present study are available upon reasonable request to the authors

